# Safety in Adolescent Behavioral Health Crisis Units: A Qualitative Analysis of Clinicians Versus Designers’ Perspectives

**DOI:** 10.64898/2026.01.28.26345086

**Authors:** Roxana Jafarifiroozabadi, Hani Patel, Paul Thomas Clements

**Author notes:** **Corresponding Author Roxana Jafarifiroozabadi, Ph.D., EDAC, Assoc. AIA** Ronald L. Skaggs, FAIA Endowed Professor in Health Facilities Design; Assistant Professor in Architecture, Texas A&M University, Department of Architecture Langford A-428, 3167 TAMU, College Station, TX 77843-3167, **E:**.

## Abstract

Safety is a foundational concern in adolescent behavioral health crisis units (BHCUs), where therapeutic care must be delivered in complex, rapidly evolving environments. However, limited research has explored how key personnel involved in shaping the environment of care in such units, such as clinicians and healthcare designers, understand and prioritize safety. To address this gap, one-hour, online semi-structured interviews were conducted with a panel of experts (N = 13) at a national level in the U.S., comprising of eight designers (healthcare designers and medical planners) and five clinicians (psychologists and psychiatric nurses) actively involved in designing or construction of BHCUs or providing care in these units for adolescent patients in the past five years. The interviews were recorded, transcribed verbatim, and analyzed via MAXQDA (2024) for qualitative content analysis. Analysis of interviews revealed 592 codes forming four preliminary categories related to safety in adolescent BHCUs: 1) Barriers and facilitators to patient safety and comfort (f = 52%), 2) Care processes and clinical workflows (f = 21%), 3) Care outcomes (f = 19%), and 4) Laws, regulations, and guidelines (f = 7%). Findings highlighted several points of divergence in clinicians versus designers’ perception of safety related to environmental features, such as nursing station designs, patients’ access and control over unit features, and furniture type or layout in the unit. Results also showed differences in understanding care processes and outcomes related to safety among the two groups. Addressing such discrepancies can contribute to the development of safer BHCUs that support adolescents’ healing.

## 1. Introduction

Safety is the paramount concern in the mental and behavioral health settings (No & Lee, 2024; Vogt et al., 2025) as incidents of aggression, violence, self-harm, or suicide can compromise safety or cause emotional distress among patients and staff (Dalton et al., 2023; Mullan et al., 2023; Thibaut et al., 2019). Despite the centrality of safety in mental and behavioral care settings, safety perception often varies across occupants and stakeholder groups. The existing literature shows discrepancies in safety perceptions between mental and behavioral health patients (Berzins et al., 2018, 2020) and stakeholders responsible for facility operations and care delivery with a focus on organizational factors and mental health service delivery (Albutt et al., 2021). However, there has been limited focus on how the physical environment of care can influence safety perceptions across the diverse groups, including healthcare designers versus those involved in management or care delivery in mental and behavioral healthcare units. Studies suggest that clinicians, who directly interact with patients and manage risk in real time, often conceptualize safety in terms of patient behaviors, staff vigilance, and crisis management (Latimer et al., 2014; Wolff et al., 2023; Wong et al., 2015) while designers optimize safety in the environment of care through improving spatial layouts, material choices, or furniture configurations (Fornefeld et al., 2024). These discrepancies in safety perceptions can become even more pronounced when serving specific patient populations, such as adolescents, who require specialized care in environments tailored to their unique needs (Connellan et al., 2013; Saxon, 2018). There is little evidence on improving safety features in adolescent mental and behavioral care environments, such as behavioral health crisis units (BHCUs), and current care models and environments are mostly adult-centric (Kim et al., 2022; Zeller, n.d.; Zeller et al., 2014a, 2014b, Fornefeld et al., 2024). To date, studies have investigated a myriad of environmental features to optimize the mental and behavioral healthcare environments, such as ligature-resistant features (Hutton et al., 2021), nursing station designs (Kalantari & Snell, 2017; Novotná et al., 2011; Rodríguez-Labajos et al., 2024; Wolff et al., 2023), seating arrangements (Shepley et al., 2016), interior detail or furnishes (Karlin & Zeiss, 2006; Pawlaczyk-Szymańska et al., 2025; Shepley et al., 2017), privacy (Novotná et al., 2011), art work or sources of positive distraction (Eisen et al., 2008; Kalantari & Snell, 2017), access to nature (Fornefeld et al., 2024; Fricke et al., 2019), lighting (Norouzi et al., 2023), comfort and autonomy (Fricke et al., 2019; Norouzi et al., 2023; Trzpuc et al., 2016), or noise (Connellan et al., 2013). Nevertheless, there is a lack of research examining how safety is prioritized, understood, and interpreted by clinicians compared to designers, as well as how these environmental features shape safety perceptions among these two groups within adolescent mental and behavioral healthcare environments. Also, very few studies have investigated safety in milieu environments (Curtis et al., 2007; Shepley et al., 2017; Trzpuc et al., 2016) accommodating selected groups of patients for receiving therapeutic interventions when experiencing mental and behavioral health crises, and little is known about the optimal layout, design features, and furniture layouts in these environments.

This study addresses this gap through a qualitative analysis of clinicians’ and designers’ perspectives on care processes and safety in adolescent mental and behavioral care environments with a focus on the milieu environment of BHCUs. By comparing how each group prioritizes and operationalizes safety, study aimed to identify alignments and discrepancies in perception of care processes, environment, and safety for adolescents experiencing mental and behavioral health crises.

## 2. Methods

This qualitative study employed online semi-structured interviews with 13 experts to capture their feedback regarding safety in adolescent BHCU environments and explore nuanced perceptions and reflections that may not be accessible through quantitative methodologies. Purposive sampling was used to ensure diversity in role, experience, and professional background of experts, including five clinicians (psychologists and psychiatric nurses) and eight designers (medical planners and healthcare designers). The inclusion criteria included being at least 18 years of age and have been actively involved in providing care or designing environments for adolescent patients experiencing behavioral health crises in the past 12 months.

All participants were provided with information sheets and consent forms prior to data collection via email. Interviews were conducted via a secure video conferencing platform (Zoom) using a laptop in a lab space on a university campus in the southern United States. Each interview lasted approximately 60 minutes and was audio and video-recorded with consent. The recorded interviews were transcribed verbatim using Zoom’s AI Companion feature. Each AI-generated interview transcript was cross-checked against the audio recordings by two trained research assistants to identify and correct any potential misalignments. Ethical approval was obtained from the Texas A&M University Institutional Review Board (IRB). Participants provided written informed consent and were assured of confidentiality and their right to withdraw. Transcripts were de-identified, and all data were stored securely in compliance with institutional guidelines.

### 2.1 Interview Questions

Interview questions and guidelines were developed separately for clinicians and designers through team discussions and a review of relevant literature. The interview questions were focused on care environments, processes, and protocols for units accommodating adolescent patients (12 – 18 years of age) with multiple co-occurring conditions, such as mental health and substance abuse issues, across varying acuity levels (high, medium, and low risk of harm to oneself or others). Additionally, the interviewees were asked to describe examples or scenarios of challenging situations (e.g., violent incidents) they had encountered in BHCUs and milieu environments and elaborate on barriers and facilitators related to the care workflows. To gather further feedback on unit layout, furniture types, and arrangement, sample images of a unit featuring different furniture were shared during the interviews, and participants’ responses were collected.

### 2.2 Data Analysis

Each interview lasted approximately 60 minutes, was audio recorded, and transcribed verbatim for analysis using MAXQDA (2024) to facilitate data management. Transcripts were analyzed using content analysis based on the methodology described by Burnard (1991). The analytic process included multiple readings of each transcript, open coding, clustering of codes into categories, and abstraction into higher-order themes. Each transcript was coded independently by at least two members of the research team. Discrepancies were discussed and resolved through consensus. Additionally, Intercoder Agreement was investigated through percentage agreement feature in MAXQDA.

## 3. Results

### 3.1 Participant Data

The interviewees (*N* = 13) included eight senior designers—comprising healthcare designers, medical planners, and behavioral health practice leaders—and five clinicians, including psychologists and psychiatric nurses. Of the 13 participants, eight were female (four designers and four clinicians) and five were male (four designers and one clinician). All participants had at least five years of experience in either designing healthcare facilities or providing care for patients with mental and behavioral health needs across the U.S. Participants were also actively involved in projects or care activities in various states across the United States, including Texas, California, North Carolina, Virginia, and New York. All participants possessed a master’s degree or higher.

### 3.2 Semi-structured Interviews: Emerging Themes and Categories

Codes (N = 592) were compared within and across interviews, with conceptually similar codes clustered together to rebuild the data into the following preliminary categories related to adolescent BHCUs: 1) Barriers and facilitators patient safety (f = 52%), 2) Care processes and clinical workflows (f = 21%), 3) Care outcomes (f = 19%), and 4) Laws, regulations, and guidelines governing the provision of mental healthcare (f = 7%). Categories and code frequencies are illustrated in Figure 1. Each category is discussed in detail in the following sections.

**Figure 1.**
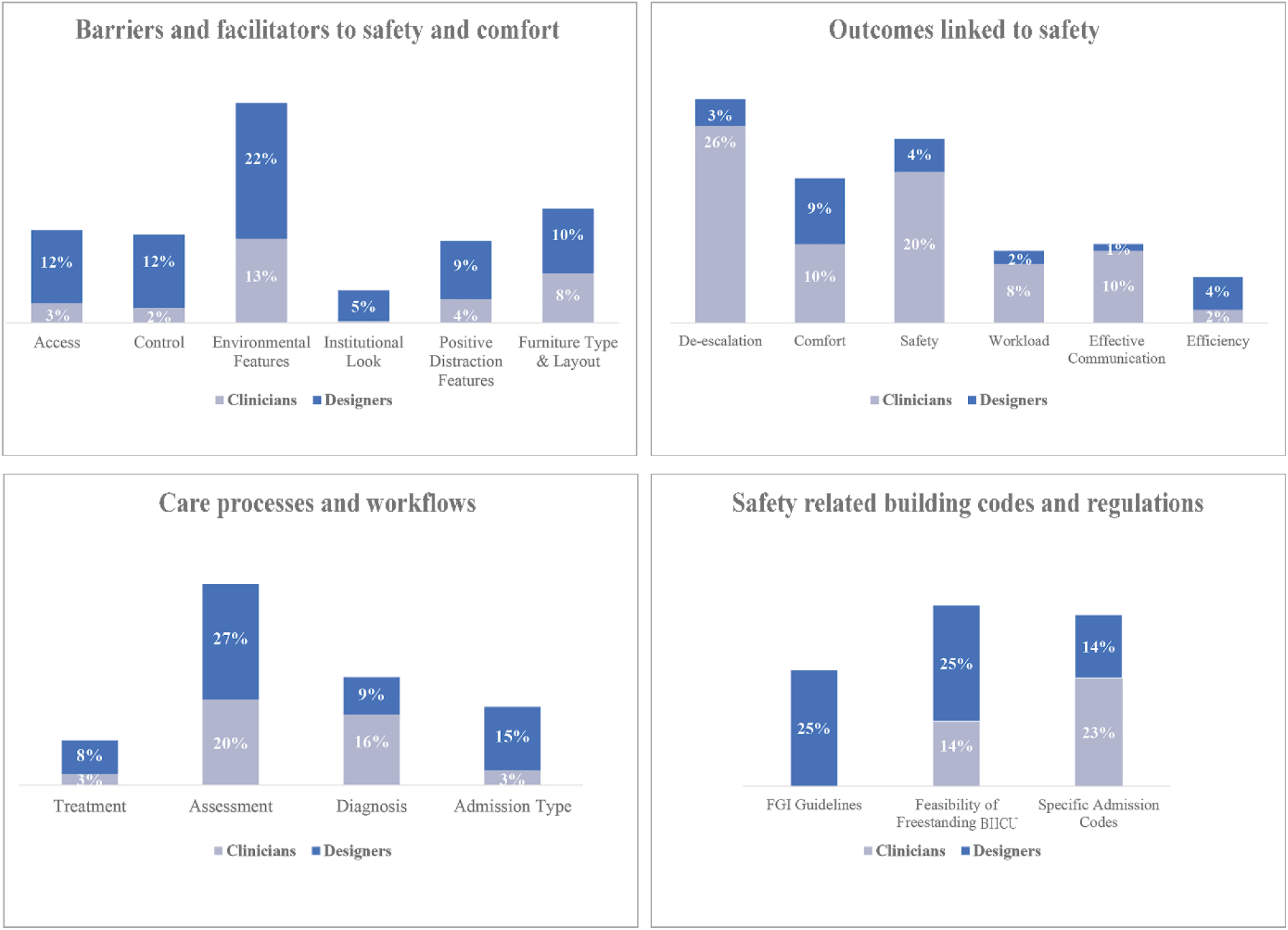
– Categories and code frequencies obtained from analysis of interviews.

#### 3.2.1 Barriers and facilitators to safety and comfort

Overall, more than half of the identified codes based on the discussions with both clinicians and designers were related to safety and comfort during and following patient admission to the facility. Factors impacting safety were categorized as environmental features, access, choice and control, and factors related to comfort included access to sources of positive distraction and lack of an institutional look within the facility.

*Safety and Environmental features:* When discussing facilitators and barriers to safety in adolescent CSU environments, the majority of identified codes related to the factors impacting safety and comfort (*f* = 35%) were focused on the environmental features withing the facility (Figure 1). Safety concerns related to the environment within the facility were mostly related to materials and items (*f* = 35%), visibility (*f* = 27%), and seclusion or separation of pathways (f = 27%), followed by comments related to safety alerts (*f* = 5%), ligature risks (*f* = 4%), and elopement risks (*f* = 3%). In discussion of materials and items, both clinicians and designers discussed safe shape and type of furniture, desks or counters in the nursing station as well as finishings and materials on the ceilings, walls, and floors (Table 1). Visibility, patient monitoring, and safety alerts were also deemed significant in the units. Both designers and clinicians indicated that it was essential to integrate the caregivers’ workstations in the milieu environment, orient caregivers ‘workstations and seats towards the patients, and ensure that all patients were always visible to the caregivers in units. Interviewees also discussed the design of caregiver workstations for optimal visibility and supervision. One clinician mentioned: “*in these units, we have a centralized location for the nurses to be able to see the milieu either through like direct line of sight or through digital devices. It’s kind of like an octopus, with a central hub in the middle and the allotment areas branching out from it”.* One designer stated:” *this (supervision) can be effectively managed from an open nursing station as opposed to a fully enclosed one with glazing”.* Another clinician added: “*I have heard from kids like they don’t like feeling like they’re in a fishbowl.”*

**Table 1.** – Clinicians versus designers’ feedback regarding the impact of interior materials and features on safety in adolescent BHCUs.

| Interior Materials, Items, and Ligature Risk | Interviewee | Comments |
| --- | --- | --- |
| Nurses’ Desk or Counter | Clinician | "When I was a nurse manager, the most damage was related to weapons of opportunity. I used to tell the nurses to stop putting things at the nursing station like scissors and purses or anything that could not be tied down. A phone and a cord were the worst injury producing thing." |
|  | Designer | "Maybe we think about how that desk in the nursing station can be designed to be a little taller, or maybe a little deeper than a typical desk. So, if somebody does come to up to the desk, they're able to do so. But if somebody's coming up to the desk, because they'd like to attack or do something negative, it's going to be much harder for them to do that." |
| Ceiling Finishes and Materials | Designer | "We could consider a secured ceiling versus standard acoustical ceiling tiles! We make sure that a ceiling is high enough that somebody couldn't get on a chair and grab a standard, you know, grid from an acoustical ceiling tile and use that as a weapon or use it (the ceiling tile) as a place where they could hide some kind of contraband or drug." |
| Wall Finishes and Materials | Designer | *"We've had patients who try to climb exterior walls in the courtyard or the building itself. So, we've talked a lot about the use of materials and making sure that we don't have materials that are too easy to climb."* |
|  | Clinician | *"Smooth finishes are probably good in case you have folks who like to, you know, hit their heads on the wall. Things that aren't incredibly textured, from a safety perspective, with calming colors are usually nice."* |
| Floor Finishes and Materials | Clinician | *"We could have some sort of squishy playground that was safe for the younger kids where they could just, you know, get their energy out"* |
| Furniture Type and Arrangement | Designer | *" We could create little nooks or seating or benches within the space that a client (patient) can go to and sort of be alone, but still in the visual, still part of the larger group."* |
|  | Clinician | *"The Ottomans, if used in the space, need to be both movable but not easy to pick up. Anything that is easy to pick up can be thrown. soft things, decorations, and pillows are great!"* |
|  | Clinician | *"The chairs should be filled with sand and should not have really sharp edges so that we can still slide them around and rearrange them for group therapy settings."* |
| Wall-mounted Items | Clinician | *"We have to make sure the pictures on the wall are screwed to the wall and cannot be removed. We have to ensure that they have plastic instead of glass and the frames are absolutely not possible to be removed or broken."* |
|  | Clinician | *"If there are TVs, we make sure that those are either high enough where folks can't get to them or placed behind plexiglass."* |
| Bathroom Fixtures | Clinician | *"We need to sure that the seclusion room toilet is a non-horseshoe shaped toilet seat... we had a patient who knelt with a towel, placed their neck in the opening of a horseshoe-, and tragically died by suicide."* |

In addition, interviewees also revealed safety concerns regarding managing agitated patients in the unit environment in crisis situations. One designer mentioned: “*I think that the biggest challenge is what happens when a patient gets agitated and disruptive to others. One solution is to have those opportunities where they can have some escape space where they can get away safely and comfortably. Hopefully not, you know, escorted to seclusion rooms, but guided to a calming space until they’re able to calm down.”* Another designer added: *“Oftentimes, we have seclusion suites at that entry point, so that if somebody is really needs to calm down or needs some extra help, that’s close to the entry point.” Also,* both interviewees emphasized the importance of separated entry points, sally ports, or waiting areas for patients who are detained under law enforcement from walk-in patients as well as visual and acoustical separation among spaced allocated to pediatric, adolescent, and adult patients. One designer mentioned: “*In standalone crisis centers, they (patients) could come into a joint lobby or dedicated entrance, and they sort of bifurcate into urgent mental health care for adolescents versus adult, and they might have their own separate waiting area, reception, and consult areas*.”

**Access:** Immediate access or lack of access to certain areas and features were deemed essential by clinicians (*f* = 3%) and designers (*f* = 12%) to ensure safety among adolescents experiencing mental and behavioral health crises. According to the designers, it is crucial for the caregivers to have immediate access to patient areas from the nursing station, medication room, documentation or teaming spaces, conference room or consultation room, and therefore adjacencies of those spaces could facilitate safety and care workflows. However, direct visibility or immediate access to these areas from patient areas are not desired. Clinicians indicated a need for immediate access to systems for alerting other providers and staff members at the time of emergency in the unit.

Both designers and clinicians mentioned that patients need to have access to storage for belongings such as cell phones and cloths, sleeping rooms or private areas, and bathrooms for showering and changing of clothes if need be. One designer stated: “*Once you receive a patient, there’s spaces for decontamination showering, you know, if they’re coming directly off of the street, or there’s been something where they’re soiled, then you may take them to a space to disrobe and clean and provide shower facilities and then fresh clothing for them. And that may happen before you do anything else, in order to get somebody where they need to be to receive that initial consultation and assessment*”. Access to nourishment was also deemed important for patients; however, designers and clinicians indicated contradicting outcomes regarding direct access to certain features in nourishment areas, such as hot beverages: “*a nourishment area can have snacks and drinks like coffee or access to ice or something…but there have been facilities that had to stop having that, because, you know, if you had the coffee and somebody wanted to hurt somebody else…”.* One clinician mentioned: “*There should be a locked kitchen where all this stuff can go in there. If you think about it, like I know for myself, it’s very frustrating to not be able to get your own food and drink what you want when you want it right*”.

**Control and choice:** Designers (*f* = 12%) and clinicians (*f* = 2%) suggested that the environment of care should provide adolescent patients with meaningful autonomy and control, allowing them to choose how and when to engage with others, regulate their level of social interaction, and access a variety of spaces that support different activities and therapeutic needs. Designers mentioned: “ *So, part of the trauma-informed voice and choice is that they (patients) can then choose to go in different spaces for different things, like, if there’s a particularly loud day room with the TV and the video gaming system that the kids are playing at, and you know, someone kind of just wants to sit in a corner, they can go to the quiet room*” or “*if there’s a rocker in the corner of the room that they (patients) can just need a moment to themselves, and you know what I mean, it’s really about giving that choice*”. Additionally, designers recommended that it is important to create sensory environments and calming rooms for patients where they can control and adjust lighting levels, lighting colors, access to natural lighting, temperature levels, and furniture arrangements in their environment: “*when we design sensory spaces, we often think about how it can be a dual purpose there…Even if it’s not a full sensory space where, you know, materials and lighting change, it’s just a space where you can do a little aromatherapy, you can have the lights dim, maybe you can have a little bit of music, and it’s just a quiet room*” or “ *I think variety is a good thing. With the seating arrangements, they can choose to recline. We also build in a lot of bench seating and window seats and other things where you can kind of experience space from different perspectives*”. Clinicians also stated the importance of having a choice over the type of snack, food, and beverages for adolescent patients: “*we had a snack cart that would come around once in the morning and the afternoon. And that just felt like a special fun time, like, oh, we can choose our own cookie, chips, and drink, and that just made it more fun and give them a little bit more sense of control without making it a free for all, like a vending machine so they could get to choose (the snacks) from*”.

**Institutional look or features:** Clinicians and designers’ comments regarding the institutional look of the facilities were mostly focused on the design of the nursing stations (enclosed versus open) within the units during patient observation and care processes. One designer mentioned: “*they (caregivers) are used to managing units from behind ten-foot polycarbonate glass that is one inch thick. That says one thing! It says I’m not approachable. You know, like, you stay there. I stay here. It starts to look very institutional, prison like, and some of the imagery coming from the institutions, let’s say crisis units, looks like a slight modification of a prison*”. While interviewees overall emphasized the removal of glass barriers between caregivers and patients to facilitate better communication and home-like feeling at their units, they pointed out safety challenges related to a more open design for caregivers’ workstations. For instance, one designer stated: “*It was very important to them (caregivers) to not feel like they were behind glass walls for their clients (patients). But as soon as the facility opened, clients were reaching over and picking up computers and throwing them. So, we’re putting up glass walls now.* “Moreover, clinicians commented on the lighting and color features in facilities for a calming and therapeutic impact on patients. One clinician mentioned: “*there should be no bright fluorescent lighting, things that make them feel like they’re interrogated.”*

**Sources of positive distraction:** The desired sources of positive distraction for adolescent patients were mostly focused on the sensory features related to lighting, tactile, acoustics, olfactory features. Tunable and color artificial lighting for the purpose of light therapy and creating calming and soothing environments were deemed highly preferred in the units. Clinicians and designers also emphasized soothing music, calming colors, and improved access to natural lighting and views of nature for adolescent patient. Designers suggested that even in environments, such as seclusion rooms, where implementation of windows could be challenging, a skylight providing natural lighting for patients could improve play an important role in improving their comfort and health. Integrating TVs and gaming devices were also debated to be essential but challenging features to be included in these units. One designer stated: “*in observation lounges (milieus) TVs are often the source of conflict in a number of ways, so that can be very challenging!”* One potential solution suggested by clinicians and designers was to provide cubicles with personal entertainment systems or devices in the milieu environment to allow for control and choice over activities and minimizing potential conflicts in the unit.

**Furniture type and layout:** In addition to discussions pertaining to safe furniture type and arrangements in previous sections, interviewees were provided with images of a proposed furniture layout for an adolescent BHCU during its construction phase, and were asked to provide feedback on the concept of color-coded, assigned seats or sleeping surfaces in the unit to personalize the shared space in the milieu and help patients understand their role in the built environment (Figure 2). Findings indicated discrepancies in clinicians versus designers’ feedback regarding effective incorporation of the furniture layout or color-coding concept into the milieu environment for adolescents experiencing mental and behavioral health crises.

**Figure 2.**
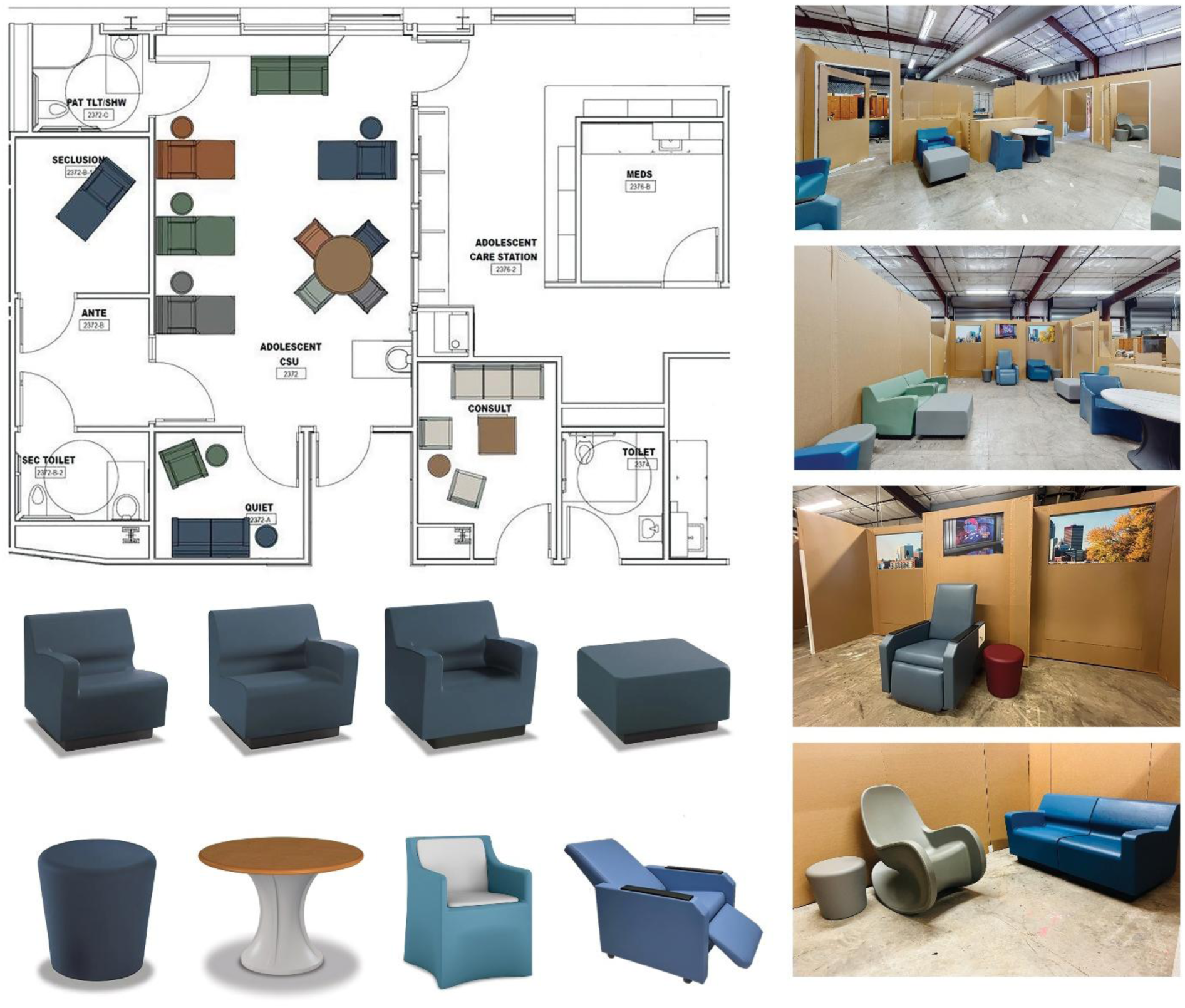
– Proposed furnished design for an adolescent Behavioral Health Crisis Unit during the pre-construction design phase.

Most designers (*n* = 6) evaluated the concept positively and considered it “an *interesting concept that merits further exploration*”. Only one designer mentioned that the concept of color-coded furniture “might not be a good idea”. Additionally, some designers highlighted lack of specific furniture types in the proposed layout, including recliners for sleeping, rockers, and bean bags. On the other hand, clinicians (*n* = 4) mostly mentioned that the concept of color-coded furniture is “not a good idea” as it may cause conflict or power struggles. They also mentioned that the concept will limit the adolescent’s choices and negatively impact care processes through increasing caregivers’ workload. Other clinicians asked at what point during care processes seat assignments should take place. Selected comments by designers and clinicians regarding the proposed furniture layout is presented in Table 2.

**Table 2.** – Clinicians versus designers’ comments on furniture Layout, color-coded furniture pieces, and assigned seating concepts in adolescent BHCUs.

| Designers | Clinicians |
| --- | --- |
| *When we look at where incidents had occurred, either between patients or between patients and staff, they were in those ambiguous spaces where it wasn't clear who owned the space. This is interesting because I feel like... If I go in there... I know that blue seat at that table is mine. Then there's the potential that everybody can just be comfortable in their own space and have fewer conflicts.* | *First thing I'm going to tell you is you (caregivers) are going to get into power struggles instantly with assigned seating. The minute you tell a teenager that they can't sit somewhere, we are going to have caregivers be spending all their time telling the teenager to get out of that chair and move into another chair. I am not dealing with this!* |
| *I think it can guide younger people, right? I think as an adult, it might feel a little constraining and it sort of takes away some of that voice and choice that we really want to give to people within the facility.”* | *My first impression honestly, was no! I think they (patients) would perceive this as control. You (caregiver) tell them you can only sit in the orange... it could also even precipitate fights.* |
| *As someone who's slightly colorblind, it really depends on their sensory perception. I think it's an interesting organizational structure. It might be fascinating to kind of explore, maybe how that is beneficial or not detrimental. We've used motifs, and I mean.* | *My most immediate thought in terms of challenges is just at which point do you assign the seat. If you assign it at the check-in, you may not yet know what those interpersonal dynamics may be. A lot of these individuals personality characteristics that make them less friendly or interpersonally engaging or overly engaging. So, I would just be concerned about that you wouldn't be able to control for that.* |
| *So, my initial reaction to the color-coded thing is that's not a good idea because it could just cause problems. You don't need to have this if somebody's already in crisis and agitated.* | *You want to give them as much choice as possible, because most of our adolescents are struggling because of the lack of choice. I think that might limit their choices or their freedom. I don't know that you need the colors.* |
| *I see that possibly being kind of like a good thing for the lounge chairs. So, for the big ones. But when it comes to the small seats around the table. I'm not sure about that!* | *That's a really great idea. I like that it kind of gives them their own space.* |
| *So, I'll start with the color coding. I think it's an interesting concept. It shows a sense of ownership. Patients an understanding of boundaries, physical boundaries. Considering that we're focusing on the adolescent population... Well, some of them may not like the color that they have, and so that could create a bit of agitation.* | *You have to remember that some of these people have delusions going on like they might fear the color orange, or they may only prefer blue if they have the obsessive-compulsive disorder (OCD).* |

#### 3.2.2 Care processes and workflows

Clinicians and designers both discussed workflows and care processes associated with safety for caregivers and adolescent patients during admission to these units. The key categories identified based on the codes extracted from both designers and clinicians’ comments included patient admission type (*f* = 18%), assessment (*f* = 47%), diagnosis (*f* = 25%), and treatment plans (*f* = 11%). Interviewees indicated that patients in these units are admitted either voluntarily, with minimum needs for de-escalation, or involuntarily requiring being transferred to the unit by the ambulance law enforcement. A designer mentioned: “*if you’ve got walk-in patients and potential law enforcement or ambulances coming in, you really want to separate those entry points. Oftentimes, those that are coming in by law enforcement may not want to be there and you want to kind of provide that dignity for their entrance through what is called a vehicle sallyport*.” A clinician added: “*They (patients) may be searched to make sure they don’t have any means on themselves to engage in any kind of harm behaviors either to themselves or others*”. In most cases, patients arriving through the sallyport are not restrained and will go through the registration and triage process for medical and/or mental health examination in a secure intake or triage space. According to the clinicians, patients’ medical records will be reviewed, and they will be interviewed and examined in a private consultation room for externalization or explosive behavior, hyper motoric indicators, nervousness, or suicidal tendencies which may pose safety risks in the unit. Upon noticing signs of aggressive behavior or active substance use, patients might be considered for other types of care or transfer to an inpatient facility. A clinician mentioned: “*we triage them (patients) into high, medium, or low acuity. Also, I would be assessing whether they have ideation, intent, and plans, and the extent to which they have intend to act on those plans. They have access to the means to carry out those plans, what access to means they have at home to determine if they need to be fully hospitalized.*” Designers added that following initial assessments, patients are guided to spaces for decontamination or showering if need be or may wait in waiting areas. One designer highlighted inclusion of seclusion areas at the time of patient intake: “*we want to also think about having a seclusion room nearby if a patient is highly agitated, and needs to be moved to a different space, temporarily, until they have calmed down enough and reached a state where they can actually participate in the intake process.”* In case patients are assessed to be in an acute crisis phase, they will not be directly transferred to the group therapy in a milieu that day until stabilized. Instead, they will be sent to the consultation room for a one-on-one treatment in a more traditional crisis stabilization unit or transferred to an inpatient facility. Following initial assessments, the treatment plan begins within the first 24 hours without getting into deep therapy.

#### 3.2.3 Outcomes linked to safety

Interviewees described the important care outcomes associated with adolescent BHCUs (Table XX). De-escalation, safety (for patient and caregiver), and comfort were the top three desired outcomes identified by both clinicians and designers for adolescent patients BHCUs followed by effective communication, reduced workload, and efficiency for caregivers. When discussing de-escalation strategies, clinicians (*f* = 26%) mostly discussed care interventions or training programs, such as PIAB (Patient-Initiated Brief Admission), PMAB (Prevention and Management of Aggressive Behavior), Crisis Prevention Institute (CPI) training, or cognitive behavioral interventions. Other strategies included incorporation of grounding techniques or separating the agitated individual from the rest of the group by sending the non-agitated patients to a different area temporarily to de-escalate or mitigating situations. Strategies, including restraining patients, assignment to seclusion rooms, or using PRN psychotropic medications (medications given with the aim of changing the patient’s mental state) were considered by clinicians as a last resort. On the other hand, designers (*f* = 3%) emphasized the role of the environmental features in de-escalation process, including acoustic qualities, soothing music, and openness of the environment. For achieving comfort and well-being as a desired care outcome, both clinicians and designers emphasized the importance of the trauma-informed approach and care environment. Home-like, soothing environments with calming colors and comfortable furniture as well as inclusion of social support and family support spaces for adolescent patients were deemed essential for achieving patient comfort during care.

When discussing safety as a desired care outcome, clinicians (*f* = 20%) mentioned safety training programs for ensuring both patient and caregivers’ safety during care activities or at the time of crisis. Regarding caregivers’ safety, one clinician mentioned: “*in physical altercation safety training programs like Handle with Care (HWC), they aim to make sure you can move out of the way safely, to keep yourself physically safe without hurting yourself or the patient*”. Another clinician mentioned: “*if there was ever a riot, we would all get into the chart room and then call 911 if we had to, or else we called a code from the other units*”. For patient safety, clinicians mentioned the importance of 15-minute checks on patients not just from the nurses’ workstation, but with walkthroughs in the unit to ensure that patients are safe. Others restated security checks for prevention of carrying contrabands or substances in case of care for adolescents with substance abuse disorders. A few clinicians also re-emphasized the importance of moving hazardous furniture or items, such as light chairs or furniture pieces out of the way or avoiding certain areas such as elevators or staircases when handling agitated patients in the unit. Designers (*f* = 4%), on the other hand, focused on safety outcomes related to environmental features such as availability of safety glasses (e.g., polycarbonate glass) or inclusion of at least two exit points in every patient-accessible area of the unit, allowing caregivers to leave the situation safely and comfortably. To achieve effective communication, clinicians discussed availability of team spaces, collaborative spaces, or conference rooms for discussing patient status or hand-offs privately. Regarding workload, most clinicians discussed caregiver to patient ratios in the unit which varied between 3-5 to one depending on the patients’ acuity level in the unit. To improve efficiency, one clinician mentioned: “*Most of these units are staffed with people that have dual competencies. Let’s say they come from the nursing world or medical world, and they have also acquired licensing to deal with psychiatric patients*”

#### 3.2.4 Safety related building codes and regulations

As indicated in Figure 1, emerging themes related to safety codes and regulations included building codes and guidelines by sources such as the Facility Guidelines Institute (FGI), specific patient admission codes, and other regulations such as freestanding crisis stabilization units (CSUs). *Facility Guidelines Institute (FGI):* Designers led the discussions regarding regulations and guidelines for designing and constructing these units based on the FGI guidelines. They pointed out the importance of following existing guidelines and building standards to ensure safety in adolescent BHCUs. One designer mentioned: *“the biggest step forward nationwide has been the introduction in the FGA guidelines to a section that is specifically addressing the design of crisis environment. So, this is new! It was not there before, and it now exists”.* However, designers highlighted challenges with the development and acceptance of guidelines across the U.S. and stated, *“FGA guidelines are NOT adopted by every state!”, and “When we wrote the FGI guidelines, we wrote them to be as loose as possible”, or “They (guidelines) are relatively specific about what needs to be provided. But there’s also a lot of flexibility in the types of spaces”*.

*Specific state regulations for patient admissions:* Both clinicians and designers had different perspectives on regulatory aspects of patient admission. Clinicians commented on legal frameworks on patient consent and treatment. One clinician stated: *“there are laws where we are required to treat anyone who comes in and says they’re suicidal wherever they’re at. So, you know, as a primary care doctor, we can’t say, well, we don’t do this, you need to go see a psychiatrist or a psychologist, we have to intercept them, and it doesn’t matter the age of the individual”.* Other clinicians added: *“At the age of 15, they can find themselves out of hospitalization or stabilization without notifying period”,* or “*You can treat without parent consent so police officer can bring them in. You can treat without parent consent if there is risk of harm to themselves or others, if there’s substance abuse, or if they’re being abused*”. One designer noted that *“They (BHCUs) are kind of an emerging thing that’s coming through and the licensing piece is catching up to that”*.

*Feasibility of providing care for adolescents in freestanding units such as CSUs*: Both clinicians and designer had mixed opinions on the feasibility of freestanding units. The contrast emerged in terms of the operational challenges vs the benefits of locating in the rural areas. One designer stated *that “the downside of having a standalone unit is that you don’t have the interaction of those providers that are on the units that are behavioral health specialists for peds. And they can’t come down and collaborate easily they have to travel to another location”.* Another designer noted that *“they might be in more rural or suburban settings, where they may not have a major medical center which to attach to anyways. So, it’s just kind of the nature of where they need to be to be accessible to the community”.* One clinician also explained the effectiveness of such unit: *“they work and they’re very feasible, especially in communities where there may not be like a psychiatric hospital”.* Another clinician added that having access to addition resources would strengthen feasibility concept and stated: *“having lab services around where they can get to doing tests pretty quickly, and screening for drugs and those types of things is fairly important*.

## 4. Discussion

This study highlights the multifaceted nature of safety in adolescent BHCUs and elaborates on how differently safety can be perceived, understood, and prioritized among clinicians versus designers. These complexities on safety were unfolded into four major themes: barriers and facilitators to safety and comfort, care processes and clinical workflows, care outcomes, and laws, regulations, and guidelines. Based on frequency of the codes extracted from the interviewees’ comments, both clinicians and designers significantly contributed to discussions related to each emerging theme. While the designers mostly provided comments on environment-related facilitators and barriers to safety in adolescent BHCUs or safety-related building codes and regulations, clinicians mostly elaborated on the desired outcomes in such units. When discussing clinical workflows, both clinicians and designers indicated a good level of understanding of care processes and provided comments regarding potential safety risks. However, the study revealed discrepancies in comments provided by clinicians versus designers on certain subcategories within these emerging themes:

The results related to barriers and facilitators to safety and comfort in BHCUs were categorized into six subcategories with environmental features within adolescent BHCUs accounting for the largest subcategory. Both clinicians and designers agreed on safe furniture, secured ceiling materials, smooth furnishing of wall and floor, and having a clear sightline toward patient areas. The study also depicted convergence among clinicians and designers on strategies, such as providing shower spaces for decontamination, and separating patient areas for different patient populations (e.g., adults versus adolescent care units) which aligns with the reports in existing literature (Pankey et al., 2022). In addition, this study revealed contradicting comments among clinicians and designers on open versus closed nurse station designs within BHCUs similar to the existing body of literature. In a study by Kalantari & Snell (2017), authors evaluated an open, ‘‘concierge style’’ communication center for patient-staff interactions in a mental health facility. While some staff favored this design strategy for its potential to enhance interactions, others expressed a more cautious “wait-and-see” stance, noting that additional evidence—particularly from longitudinal use and across different patient populations—would be needed to determine whether the design introduces safety risks. Another study in an inpatient psychiatric unit indicated similar notions with open station facilitating connection with patients while staff experienced interruptions or raised concern about confidentiality and safety in the unit (Shattell et al., 2015).

Other subcategories explored in this theme included comments regarding access to key care areas for staff or patient access and control for features, such as nourishments. While designers prioritized staff access to medication rooms, nursing stations, and other staff areas facilitating their workflow, clinicians emphasized features, such as the need for immediate access to alert systems in high-risk scenarios. Another discrepancy emerged regarding patient access to nourishments within adolescent BHCUs, with designers expressing safety concerns related to potential self-harm or harm to others from certain items, such as hot coffee. The existing research also emphasizes the importance of choice and control over features in mental and behavioral health environments, such as music panels, lighting, colors, and acoustics (Curtis et al., 2007; Hutton et al., 2021; Norouzi et al., 2023; Trzpuc et al., 2016). One study even showed that adolescent patients felt contained when they did not have access to what they needed during their stay in units (Hutton et al., 2021). These findings highlight the need to balance access to features, such as nourishments in adolescent BHCUs through creative solutions and coordinated strategies involving designers, clinicians, and stakeholders, highlighting the need for further research focused on this patient population.

Also, clinicians and designers in this study emphasized that incorporation of positive distraction sources, such as calming colors and lighting, tactile, music, olfactory, access to views of nature would benefit patients and reduce agitation in such units as evidenced by the existing literature (Eisen et al., 2008; Norouzi et al., 2023; Ulrich et al., 2018). While this study revealed contrasting opinions regarding access to TV and gaming devices as a positive distraction versus a source of conflict in adolescent BHCUs among clinicians and designers, existing literature supports that TV and mobile devices can act as a distraction to reduce the risk of conflict (Connellan et al., 2013). Nevertheless, designers in the present study provided design strategies through incorporating cubicles in units with controlled access to personal entertainment to help balance patient autonomy while reducing the risk of conflict or acoustic disturbance for others.

This study further explored unique strategies for furniture type and layout in adolescent milieu environments in BHCUs, including inclusion of color-coded, assigned seating options to the unit. While designers were open to further explore this strategy, some clinicians showed strong concerns and indicated that this strategy could result in power struggles. Current literature has not explored this dimension, but it emphasizes including mixed, ligature-resistant furniture options (Shepley et al., 2016; Hutton et al., 2021) with smooth finishes that promote safety for patients in BHCUs in general (Karlin & Zeiss, 2006; Pawlaczyk-Szymańska et al., 2025; Shepley et al., 2017).

While both clinician and designer groups appeared proficient in providing detailed descriptions of care steps and processes in adolescent BHCUs, the key factors and steps in care processes were perceived differently. Designers mostly described the care workflows and assessment processes as a sequence of spaces through which adolescent patients move during their stay in the unit, including the sally port, decontamination room, seclusion room, consultation room, and milieu. On the other hand, clinician mainly elaborated on the clinical procedures and protocols, including debriefing, administrative processes, and assessment tools for diagnosis or determining patients’ acuity level. Such discrepancies can shape how each group perceives safety or strategies to address potential risks during patient journey in the unit. Existing research and safety guidelines provide recommendations, strategies and tools regarding both the environment of care and care protocols and procedures to improve safety upon patient admission to adolescent BHCUs (Pankey et al., 2022). However, it remains unclear how these recommendations and guidelines interact to improve safety in adolescent BHCUs.

Similar discrepancies were also evident in comments provided by designers versus clinicians regarding the outcomes related to safety in adolescent BHCUs. When discussing de-escalation strategies, Clinicians mostly discussed interventions and training strategies such as PIAB, PMAB, or (CPI) training which align well with the current studies underscoring restraint reduction and safety improvements in such settings (Dalton et al., 2023; Mullan et al., 2023; Thibaut et al., 2019). On the other hand, designers mainly discussed environment-related features, including soothing music, acoustic qualities, open layout considerations for patient de-escalation to promote a therapeutic environment in adolescent BHCUs (Trzpuc et al., 2016). Although all the strategies mentioned above can contribute to de-escalation of agitated behavior and mental stability for the adolescent patients, it is unclear which combination of strategies could be most helpful in achieving these outcomes. Despite the identified discrepancies in this study, both designers and clinicians agreed that inclusion of family spaces is essential for improving comfort and well-being in adolescent BHCUs. Nevertheless, reports from the existing literature remain contradicting and show that while some adolescent patients are not comfortable interacting with parents during their stay, (Higham et al., 2012) others benefited from family interventions during their recovery (Bruns & Burchard, 2000; Ginnis et al., 2015).

Findings related to codes and regulations also revealed divergent perspectives around patient admission regulations and feasibility of free-standing units. Clinicians emphasized patient consent and treatment priorities, including “no wrong door” approaches and admission without consent in acute cases among adolescents, whereas designers focused on operational and licensure challenges, noting the absence of specific regulations in some states. Both groups expressed mixed views on freestanding units, with designers citing staffing and pediatric expertise concerns and clinicians acknowledging their value in rural settings to provide safe and timely care for adolescents.

## Limitations

The study has several limitations which should be considered while interpreting findings as it primarily focused on safety in adolescent BHCUs and had a small sample size including clinicians and designers from the U.S. only. These factors could limit the generalizability of this study to BHCUs serving other patient populations or adolescent BHCUs outside of the U.S. Moreover, this study did not investigate patients or their family members’ perspectives on safety in such units. Nevertheless, the study sets the ground for understanding divergence and agreement in opinions of national experts, including clinicians and designers regarding safety in the adolescent BHCU care environments. Additional research is required to validate findings reported in this study with larger sample of national and international experts as well as adolescent patient and caregiver populations.

## 5. Conclusion

This study contributes new knowledge by revealing how clinicians and designers perceive safety in adolescent BHCUs through four analytical themes derived from content analysis of semi-structured interviews, including environmental features, workflows and care processes, outcomes, and safety codes and regulations. Findings highlighted several points of divergence and convergence in perspectives between the two groups involved in shaping the care environment in adolescent BHCUs, and helped evaluate design strategies that could improve patient and staff safety or optimize care outcomes in adolescent milieu environments. Future studies can address discrepancies in feedback regarding design features, such as nurse station designs in such units to balance safety and interaction among staff and patients. Future research should also examine innovative approaches to balancing access, autonomy, and environmental control for adolescent patients with varying levels of acuity in BHCUs, particularly within the milieu environment. Engaging a broader range of key personnel and stakeholders in these discussions may further inform the development of safer care environments that better support adolescents’ healing and comfort.

## Data Availability

All data produced in the present study are available upon reasonable request to the authors

## Acknowledgment

The authors would like to thank the Texas A&M College of Architecture, Norix, and Solacefurniture for their support for this study.

## Ethical Considerations

Ethical approval for this study was obtained from The Texas A&M University Institutional Review Board, STUDY2024-0920. All participants provided written consent before they participated in the study.

## Contribution List

**Roxana Jafarifiroozabadi:** Supervision, methodology, investigation, conceptualization, data collection, data analysis, manuscript writing and editing, ethics approval coordination.

**Hani Patel:** Data collection, Data analysis, manuscript writing, and editing.

**Paul Thomas Clements:** Manuscript writing, reviewing, and editing.

## “Not Peer-reviewed” disclaimer

This manuscript is a preprint and has not been peer-reviewed. It should not be used to guide clinical practice.

